# Bifurcation Coronary Intervention is Associated with Higher Mortality and Complications

**DOI:** 10.1101/2023.12.11.23299826

**Authors:** Allistair Nathan, Mehrtash Hashemzadeh, Mohammad Reza Movahed

## Abstract

**Background:** Percutaneous coronary intervention (PCI) in patients with bifurcation lesions is associated with higher complexity and adverse outcomes. The goal of this study was to evaluate the inpatient outcomes of patients with PCI of bifurcation lesions.

**Methods:** The National Inpatient Sample (NIS) database, years 2016-2020, was studied using ICD 10 codes. Patients with bifurcation lesion PCI were compared to other PCIs excluding chronic total occlusions (CTO). We evaluated post-procedural inpatient mortality and complications.

**Results:** PCI in patients with a bifurcation lesion was associated with higher mortality and post-procedural complications. A weighted total of 9,795,154 patients underwent PCI, with 43,480 having a bifurcation lesion. The bifurcation group had a 3.79% mortality rate vs 2.56%. (OR, 1.50; CI:1.34–1.68; p<0.001). After adjusting for age, sex, race, diabetes mellitus, and chronic kidney disease, bifurcation lesion PCI remained significantly associated with higher mortality (OR, 1.68; 95% CI, 1.49– 1.88; p<0.001). Furthermore, bifurcation PCI was associated with higher rates of myocardial infarction (OR, 2.26; 95% CI, 1.68 – 3.06; p<0.001), coronary perforation (OR, 7.97; 95% CI, 6.25-10.17; p<0.001), tamponade (OR, 3.46; 95% CI, 2.49-4.82, p<0.001), and procedural bleeding (OR, 5.71; 95% CI, 4.85-6.71, p<0.001). All post-procedural complications were more than 4 times more likely in patients with bifurcation than in those without (OR, 4.33; 95% CI, 3.83-4.88; p<0.001).

**Conclusion:** Using a large national inpatient database, PCI performed in patients with a Non-CTO bifurcation lesions were associated with significantly higher mortality and post-procedural complications.

## Introduction

A bifurcation lesion occurs when an atherosclerotic plaque develops in a coronary artery at the point at which a side branch deviates from the main vessel branch.^1^ The main branch and side branch involvement can vary, leading to many classifications of bifurcation lesions.^2^ A variety of lesion classification systems have been published.^3-7^ The Movahed classification in comparison to the Medina classification simplifies true bifurcation lesion to one simple category called B2 (B for bifurcation, 2 both ostia are involved) and also incorporates information about other anatomical or clinical features that are not present in the Medina classification. Bifurcation lesions may account for up to 15-20% of lesions undergoing percutaneous coronary intervention (PCI).^8^ The interventional complexity of bifurcation lesions is well established, and treatment has been associated with higher rates of procedural complications and vessel restenosis.^8^ Despite such risks and complexity, PCI involving bifurcation is not uncommon.^9^ Considering the complexity of PCI involving bifurcation lesions, the goal of our study was to evaluate complications and mortality associated with bifurcation PCI using the largest available inpatient database.

## Methods

### Data Source

This study’s methodology was largely derived from that of the previous study conducted by Nathan et al, which considered percutaneous coronary intervention in patients with chronic total occlusion.^10^ This study’s patient cohort was generated using the National Inpatient Sample (NIS), Healthcare Cost and Utilization Project (HCUP), and Agency for Healthcare Research and Quality. The NIS database is representative of 98% of the total United States population, as it weights discharge data of about 35 million patients.^11^ NIS HCUP data is publicly available and is deidentified, thus the study was exempt from institutional review board approval.

### Study Population

The NIS database years 2016-2020 were considered when generating the study population. The NIS database was queried, and the study population was generated, using both International Classification of Diseases, Tenth Revision, Clinical Modification (ICD-10-CM) as well as International Classification of Diseases, Tenth Revision, Procedure Coding System (ICD-10-PCS) codes. As in the previous study conducted by Nathan et al., patients having undergone percutaneous coronary intervention were identified using the ICD-10-PCS codes 02703(4-7)Z, 02703(D-G)Z, 02703TZ, 02713(4-7)Z, 02713(D-G)Z, 02713TZ, 02723(4-7)Z, 02723(D-G)Z, 02723TZ, 02733(4-7)Z, 02733(D-G)Z, 02733TZ, 02H(0-3)3DZ, 02H(0-3)3YZ, 027(0-3)3ZZ, 02C(0-3)3Z7, 02C(0-3)3ZZ, 02F(0-3)3ZZ. This study cohort was further stratified using the ICD-10-PCS codes 02703(4-7)6, 02703(D-G,T,Z)6, 02713(4-7)6, 02713(D-G,T,Z)6, 02723(4-7)6, 02723(D-G,T,Z)6, 02733(4-7)6, 02733(D-G,T,Z)6, and 02C(0-3)3Z6 to identify patients with a bifurcation lesion. Only patients older than 30 years old were considered. Furthermore, we exclude PCI performed in the setting of chronic total occlusions (CTO). Upon completion of the multivariate analysis of high-risk baseline characteristics, any characteristics that were significantly different between the two populations were added to the multivariate analysis.

### Study Outcomes

Examined patient outcomes included mortality, myocardial infarction (I97.89), contrast-induced nephropathy (N99.0), coronary perforation (I97.51), procedural bleeding (I97.410, I97.411, I97.610, I97.611, I97.630, I97.631), tamponade (I31.4), acute postprocedural respiratory failure (J95.821), postprocedural cerebrovascular infarction (I97.821), and major adverse cardiac events. Major adverse cardiac events included any previously mentioned cardiac complication, as well as mortality. The multivariate analysis of mortality adjusted for comorbidies, age, gender, and race.

### Statistical Analysis

Statistical analysis methods were drawn from the previous study conducted by Nathan et al.^10^ Patient demographic, clinical, and hospital characteristics were reported as means, with 95% confidence intervals for continuous variables and proportions, and 95% confidence intervals for categorical variables. Trend analysis over time was assessed using Chi-squared analysis for categorical outcomes and univariate linear regression for continuous variables. Multivariable logistic regression ascertained the odds of binary clinical outcomes relative to patient and hospital characteristics as well as the odds of clinical outcomes over time. All analyses were conducted following the implementation of population discharge weights. All p-values are 2-sided and p<0.05 was considered statistically significant. Data were analyzed using STATA 17 (Stata Corporation, College Station, TX).

## Results

A weighted total of 9,795,154 adult patients were identified in the NIS HCUP database who underwent percutaneous coronary intervention without chronic total occlusion from 2016-2020. Of these patients, 43,480 had a bifurcation lesion. The average patient age was 70.12 years (CI: 70.06-70.18), and more men (63.4%) underwent percutaneous coronary intervention for their bifurcation lesion than women (36.6%). Caucasian patients composed the majority of the study cohort (77.1%). These and other study cohort characteristics may be found in *Table*. Univariate analysis demonstrated that in patients treated with percutaneous coronary intervention, those with a bifurcation lesion experienced a significantly higher rate of mortality than those without. The bifurcation group had a 3.79% mortality rate vs 2.56%. excluding CTOs (odds ratio [OR], 1.50; 95% confidence interval [CI], 1.34-1.68). Procedural complications were also significantly higher in those with a bifurcation lesion. Patients with a bifurcation lesion had significantly higher rates of myocardial infarction (OR, 2.26; 95% CI, 1.68-3.06, p<0.001), perforation (OR, 7.97; 95% CI, 6.25-10.17; p<0.001), procedural bleeding (OR, 5.71; 95% CI, 4.85-6.71, p<0.001), and cardiac tamponade (OR, 3.46; 95% CI, 2.49-4.82; p<0.001) (*Figure 1*). Overall, these patients had higher rates of all post-procedural complications (OR, 4.33; 95% CI, 3.83-4.88; p<0.001) as well as major adverse cardiac events (OR, 2.16; 95% CI, 1.98-2.35; p<0.001) (*Figure*).

**Table:** Characteristic of the population:

|  | **Total** | **Bifurcation** | **Non-**<br>**Bifurcation** | **p-value** |
| --- | --- | --- | --- | --- |
| **Total Population** | 9,795,154 | 43,480 | 9,751,674 |  |
| **Age (Mean ± SD)** | 70.12±12.12 | 66.65±12.23 | 70.13±12.12 | <0.001 |
| **Gender** |  |  |  | <0.001 |
| Male | 63.43% | 68.86% | 63.40% |  |
| Female | 36.57% | 31.14% | 36.60% |  |
| **Race** |  |  |  | <0.001 |
| White | 77.07% | 76.14% | 77.08% |  |
| Black | 10.62% | 7.62% | 10.64% |  |
| Hispanic | 7.07% | 8.42% | 7.06% |  |
| Asian/Pac Isl | 2.11% | 3.20% | 2.10% |  |
| Native American | 0.51% | 0.94% | 0.51% |  |
| Others | 2.62% | 3.68% | 2.61% |  |
| **Systolic Heart Failure** | 19.66% | 24.06% | 19.65% | <0.001 |
| **PCI Three Vessel** | 0.64% | 10.17% | 0.60% | <0.001 |
| **Diabetes** | 46.41% | 41.41% | 46.44% | <0.001 |
| **Chronic Kidney Disease** | 31.40% | 23.78% | 31.43% | <0.001 |
| **Hypertension** | 87.82% | 83.13% | 87.74% | <0.001 |
| **COPD** | 26.60% | 15.34% | 26.65% | <0.001 |
| **STEMI** | 9.09% | 28.45% | 9.01% | <0.001 |
| **Non-STEMI** | 13.66% | 45.30% | 13.52% | <0.001 |
| **Prior Percutaneous**<br>**Coronary Intervention** | 81.79% | 23.03% | 82.07% | <0.001 |
| **History of Coronary Artery**<br>**Bypass Graft** | 15.83% | 7.57% | 15.86% | <0.001 |
| **History of Anemia** | 0.04% | 0.05% | 0.04% | 0.58 |
| **History of Cardiomyopathy** | 0.86% | 0.52% | 0.86% | <0.001 |
| **Previous Smokers** | 32.89% | 26.24% | 32.92% | <0.001 |
| **Current Smokers** | 14.43% | 16.90% | 14.42% | <0.001 |
| **Atrial Fibrillation/Flutter** | 27.29% | 18.14% | 27.34% | <0.001 |
| **Valvular Heart Disease** | 8.29% | 10.36% | 8.28% | <0.001 |
| **Endocarditis** | 0.09% | 0.05% | 0.09% | 0.15 |
| **History of Stroke** | 2.47% | 1.62% | 2.47% | <0.001 |
| **Angina** | 0.06% | 0.07% | 0.06% | 0.75 |

**Figure:**
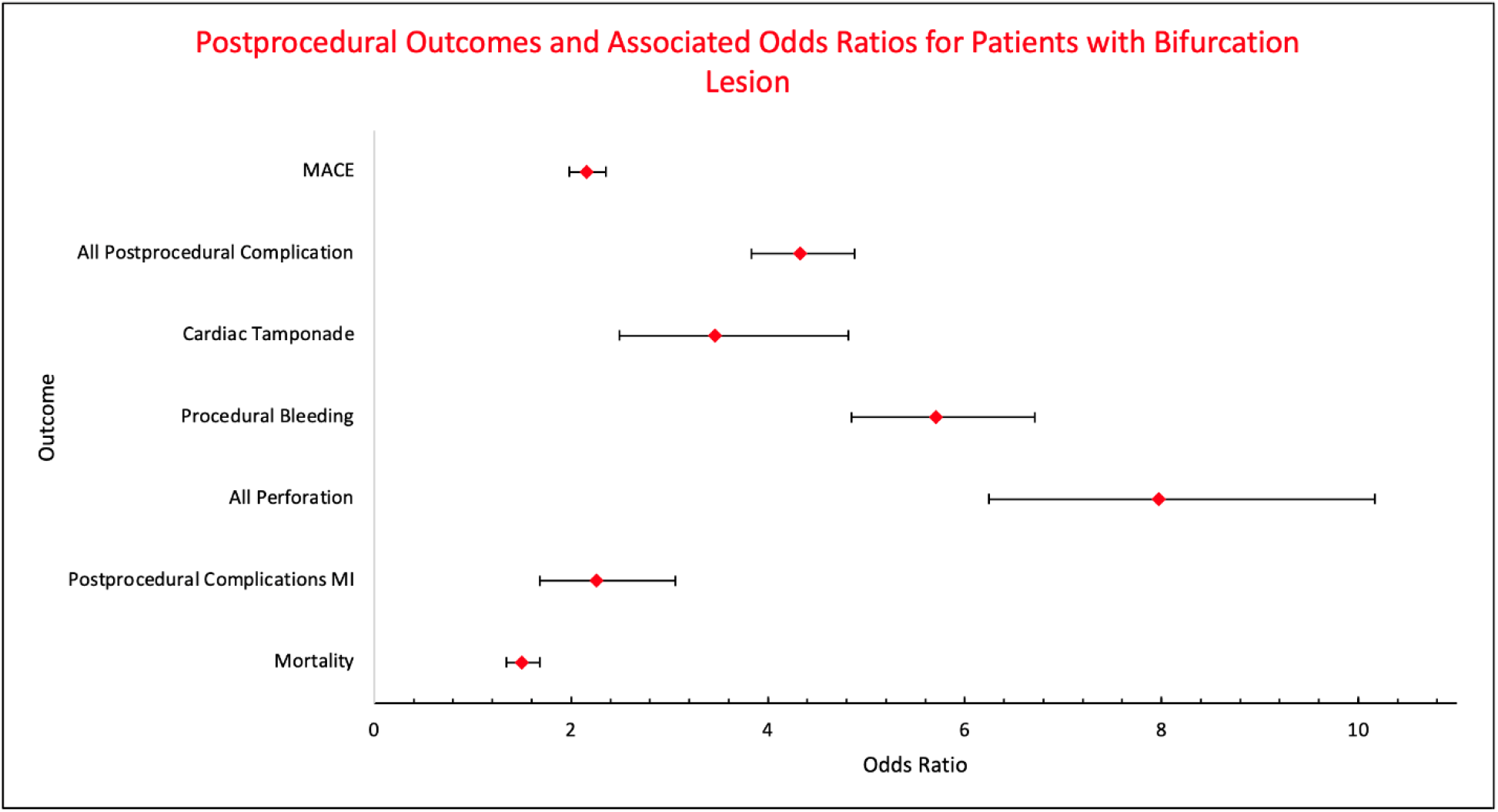
Bifurcation lesion percutaneous coronary intervention is associated with significantly increased risk of morbidity and mortality when compared to non-bifurcation lesions.

Additionally, multivariate analysis was performed adjusting for diabetes mellitus, chronic kidney disease, age, gender, and race. Upon making these adjustments, the presence of a bifurcation lesion remained associated with a significantly higher risk of mortality when treated with percutaneous coronary intervention than in those without (OR, 1.68; 95% CI, 1.49-1.88, p<0.001) (*Figure*).

## Discussion

Bifurcation lesions have been reported to be involved in up to 21% of percutaneous coronary intervention (PCI) procedures.^12^ Our study identified 43,480 bifurcation lesion interventions between 2016 and 2020. The disparity in the prevalence of bifurcation lesions reported in the literature and that found in the NIS database is likely due to the database not capturing all classifications of bifurcation lesions. The 2021 American College of Cardiology/American Heart Association Guideline for Coronary Artery Revascularization actually recommends coronary artery bypass grafting over percutaneous coronary interventions in the context of complex lesions.^13^ Additionally, Serruys et al. found that in 1,800 patients with three-vessel or left main coronary artery disease, PCI was associated with higher major adverse cardiac events and cerebrovascular events when compared with coronary artery bypass grafting.^14^

If the choice to treat a bifurcation lesion with PCI is made, there is a significantly higher risk for adverse events compared to treating a non-bifurcation lesion.^15^ This is in large part due to the increased complexity of bifurcation procedures.^16^ For example, Lam et al. found that peri-procedural myocardial infarction rates were higher in patients with bifurcation lesions than in those without, in their study investigating Resolute and Xience V stents.^17^ Additionally, a report from the SPIRIT V single-arm study found that 30-day rates of death, myocardial infarction, and target vessel revascularization were higher in patients with bifurcation lesions PCI.^18^ This increase in risk was found to persist well beyond 30 days, as Grundeken et al. found increased risk of death, myocardial infarction, and target vessel revascularization at the 5-year time point.^19^

When the results of these previous studies are compared alongside those of our study, it is clear that PCI of bifurcation lesions carries a much higher risk than non-bifurcation lesions (*Figure 1*). Our study, which is the largest retrospective analysis of PCI outcomes in patients with and without bifurcation lesions, found significantly higher rates of morbidity and complications in those with bifurcation lesions. Even after multivariate analysis, our study found that mortality was still 68% more likely in patients with bifurcation lesions. Clearly, this study and those previously mentioned studies demonstrate that the increased risk of PCI in patients with bifurcation lesions should not be underestimated.

In the specific context of bifurcation lesions, the particular arteries involved must be considered. This is well demonstrated by Kim et al., who found that only 20% of non-left main small branches supplied greater than 10% of fractional myocardial mass.^22^ Suleiman et al. specifically discussed that if only the side branch has a lesion (Movahed B1s), initial medical therapy should be first considered due to the low myocardial mass often supplied by these lesions and the risk of main vessel damage with stenting.^12^ Thus, given these findings and the increased risk of bifurcation interventional management we have demonstrated in our study, it is evident that special consideration should be given to the particular vessels involved in a bifurcation lesion and the myocardial mass supplied when considering the choice of treatment. PCI should be reserved for highly unstable lesions supplying significant portions of the myocardium.

A significant limitation of this study and all bifurcation studies are related to using media classification that separates bifurcation lesions into three unnecessary subgroups (1,1,1; 1,0,1; 0,1,1) without having any suffixes that could further characterize a specific bifurcation lesion. The Movahed bifurcation classification is the solution. First of all, it simplifies all true bifurcation lesions into one simple category called B2 lesion (B for bifurcation, 2 meaning both ostia have disease). Second of all, unlimited suffixes can be added for a given bifurcation description if needed for clinical or research purposes.^23^ Movahed et al. emphasize the importance of lesion classification when choosing an interventional approach and stent choice, highlighting that true bifurcations lesions are at the highest risk for complications whereas side-branch-only lesions (B1s may not need intervention unless the side branch is a large vessel.^4^ Thus, while our study demonstrated a very high risk associated with percutaneous coronary intervention of bifurcation lesions, the risk is probably much lower if only non-true bifurcation lesions were evaluated warranting future investigation.

### Limitations

It is important to recognize the other limitations of our study. IICD-10 coding has inherent limitations and many bifurcation lesions were most likely not coded as such. Furthermore, while we consider significant medical comorbidities, the lack of follow-up data precludes the assessment of longer-term outcomes, such as quality of life metrics. We could not separate true bifurcation lesions (Movahed B2 lesions) from others limiting our results to all bifurcation and not the high risk B2 lesions subgroup.

## Conclusion

Considering our study’s findings and those of the previous studies discussed, a significant body of evidence now highlights the complexity and risk of choosing to treat coronary bifurcation lesions with PCI. We have demonstrated that the treatment of non-CTO bifurcation lesions with percutaneous coronary intervention is associated with significantly higher morbidity and mortality than simple single-vessel PCI. Thus, the treatment of bifurcation lesions with PCI should be more selective.

## Data Availability

NIS publically available

## Conflict of interest

None

## Funding

none

## Notes

### Competing Interest Statement

The authors have declared no competing interest.

### Funding Statement

No Funding

### Author Declarations

NIS data base

## References

1. Louvard Y, Medina A. Definitions and classifications of bifurcation lesions and treatment. EuroIntervention. 2015;11 Suppl V:V23–26. doi:10.4244/EIJV11SVA5

2. Movahed MR, Stinis CT. A new proposed simplified classification of coronary artery bifurcation lesions and bifurcation interventional techniques. J Invasive Cardiol. 2006;18(5):199–204.

3. Sanborn TA. Bifurcation classification schemes: impact of lesion morphology on development of a treatment strategy. Rev Cardiovasc Med. 2010;11 Suppl 1:S11–16. doi:10.3909/ricm11S1S0001

4. Movahed MR. Coronary artery bifurcation lesion classifications, interventional techniques and clinical outcome. Expert Rev Cardiovasc Ther. 2008;6(2):261–274. doi:10.1586/14779072.6.2.261

5. Movahed MR. Quantitative angiographic methods for bifurcation lesions: A consensus statement from the European bifurcation group. Shortcoming of the Medina classification as a preferred classification for coronary artery bifurcation lesions in comparison to the Movahed classification. Catheterization and Cardiovascular Interventions. 2009;74(5):817–818. doi:10.1002/ccd.22082

6. Movahed MR. The Movahed Coronary Bifurcation Lesion Classification Introduces Limitless Optional Suffixes That Can Easily be Used for Clinical Use or Coding Purposes. Anatol J Cardiol. 2023;27(5):295–296. doi:10.14744/AnatolJCardiol.2023.3182

7. Movahed MR. What We Should Know About Bifurcation Disease. JACC: Cardiovascular Interventions. 2008;1(5):595–596. doi:10.1016/j.jcin.2008.08.003

8. Dash D. Recent perspective on coronary artery bifurcation interventions. Heart Asia. 2014;6(1):18–25. doi:10.1136/heartasia-2013-010451

9. Boden WE, O’Rourke RA, Teo KK, et al. Optimal medical therapy with or without PCI for stable coronary disease. N Engl J Med. 2007;356(15):1503–1516. doi:10.1056/NEJMoa070829

10. Nathan A, Hashemzadeh M, Movahed MR. Percutaneous Coronary Intervention of Chronic Total Occlusion Associated with Higher Inpatient Mortality and Complications Compared With Non-CTO Lesions. Am J Med. Published online June 24, 2023:S0002-9343(23)00397-2. doi:10.1016/j.amjmed.2023.06.004

11. Gacutan K. HEALTHCARE COST AND UTILIZATION PROJECT — HCUP. Published online 2020.

12. Suleiman S, Coughlan J, Touma G, Szirt R. Contemporary Management of Isolated Ostial Side Branch Disease: An Evidence-based Approach to Medina 001 Bifurcations. Interv Cardiol. 2021;16:e06. doi:10.15420/icr.2020.30

13. Lawton JS, Tamis-Holland JE, Bangalore S, et al. 2021 ACC/AHA/SCAI Guideline for Coronary Artery Revascularization. Journal of the American College of Cardiology. 2022;79(2):e21–e129. doi:10.1016/j.jacc.2021.09.006

14. Serruys PW, Morice MC, Kappetein AP, et al. Percutaneous Coronary Intervention versus Coronary-Artery Bypass Grafting for Severe Coronary Artery Disease. New England Journal of Medicine. 2009;360(10):961–972. doi:10.1056/NEJMoa0804626

15. Burzotta F, Annone U, Paraggio L, et al. Clinical outcome after percutaneous coronary intervention with drug-eluting stent in bifurcation and nonbifurcation lesions: a meta-analysis of 23 981 patients. Coronary Artery Disease. 2020;31(5):438. doi:10.1097/MCA.0000000000000847

16. Tsuchida K, Colombo A, Lefèvre T, et al. The clinical outcome of percutaneous treatment of bifurcation lesions in multivessel coronary artery disease with the sirolimus-eluting stent: insights from the Arterial Revascularization Therapies Study part II (ARTS II). European Heart Journal. 2007;28(4):433–442. doi:10.1093/eurheartj/ehl539

17. Lam MK, Sen H, van Houwelingen KG, et al. Three-year clinical outcome of patients with bifurcation treatment with second-generation Resolute and Xience V stents in the randomized TWENTE trial. Am Heart J. 2015;169(1):69–77. doi:10.1016/j.ahj.2014.10.011

18. Džavík V, Kaul U, Guagliumi G, et al. Two-year outcomes after deployment of XIENCE V everolimus-eluting stents in patients undergoing percutaneous coronary intervention of bifurcation lesions: a report from the SPIRIT V single arm study. Catheter Cardiovasc Interv. 2013;82(3):E163–172. doi:10.1002/ccd.24775

19. Grundeken MJ, Wykrzykowska JJ, Ishibashi Y, et al. First generation versus second generation drug-eluting stents for the treatment of bifurcations: 5-year follow-up of the LEADERS all-comers randomized trial. Catheter Cardiovasc Interv. 2016;87(7):E248–260. doi:10.1002/ccd.26344

20. Al-Lamee R, Thompson D, Dehbi HM, et al. Percutaneous coronary intervention in stable angina (ORBITA): a double-blind, randomised controlled trial. Lancet. 2018;391(10115):31–40. doi:10.1016/S0140-6736(17)32714-9

21. Hachamovitch R, Hayes SW, Friedman JD, Cohen I, Berman DS. Comparison of the shortterm survival benefit associated with revascularization compared with medical therapy in patients with no prior coronary artery disease undergoing stress myocardial perfusion single photon emission computed tomography. Circulation. 2003;107(23):2900–2907. doi:10.1161/01.CIR.0000072790.23090.41

22. Kim HY, Doh JH, Lim HS, et al. Identification of Coronary Artery Side Branch Supplying Myocardial Mass That May Benefit From Revascularization. JACC Cardiovasc Interv. 2017;10(6):571–581. doi:10.1016/j.jcin.2016.11.033

23. Movahed MR. Major Limitations of Randomized Clinical Trials Involving Coronary Artery Bifurcation Interventions: Time for Redesigning Clinical Trials by Involving Only True Bifurcation Lesions and Using Appropriate Bifurcation Classification. Journal of Interventional Cardiology. 2011;24(4):295–301. doi:10.1111/j.1540-8183.2011.00631.x

